# Estimated inequities in COVID-19 infection fatality rates by ethnicity for Aotearoa New Zealand

**DOI:** 10.1101/2020.04.20.20073437

**Authors:** Nicholas Steyn, Rachelle N. Binny, Kate Hannah, Shaun C. Hendy, Alex James, Tahu Kukutai, Audrey Lustig, Melissa McLeod, Michael J. Plank, Kannan Ridings, Andrew Sporle

## Abstract

There is limited evidence as to how COVID-19 infection fatality rates (IFR) may vary by ethnicity. We combine demographic and health data for ethnic groupings in Aotearoa New Zealand with international data on IFR for different age groups to estimate inequities in IFR by ethnicity. We find that, if age is the dominant factor determining IFR, estimated IFR for Māori is around 50% higher than non-Māori. If underlying health conditions are more important than age per se, then estimated IFR for Māori is more than 2.5 times that of New Zealand European, and estimated IFR for Pasifika is almost double that of New Zealand European. IFRs for Māori and Pasifika are likely to be increased above these estimates by racism within the healthcare system and other inequities not reflected in official data. IFR does not account for differences among ethnicities in COVID-19 incidence, which could be higher in Māori and Pasifika as a result of crowded housing and higher inter- generational contact rates. These factors should be included in future disease incidence modelling. The communities at the highest risk will be those with elderly populations, and Māori and Pasifika communities, where the compounded effects of underlying health conditions, socioeconomic disadvantage, and structural racism result in imbricated risk of contracting COVID-19, becoming unwell, and death.

## Introduction

The COVID-19 outbreak originated in Wuhan China in November 2019 before spreading globally to become a pandemic in March 2020. Reported fatality rates vary widely between different populations and regions from 0.25% to over 3% (CDC, 2020; China CDC, 2020; Ferguson 2020; Mizumoto 2020; Wilson 2020; World Health Organisation, 2020). At least two different measures of fatality rate exist: the infection fatality rate (IFR, ratio of fatalities to total infections) and the case fatality rate (CFR, ratio of fatalities to clinical cases). The IFR can be significantly lower than the CFR if there is a large proportion of subclinical infections. Obtaining accurate estimates of both the CFR or IFR can be difficult, particularly in the early stages of an epidemic. One reason for this is the difficulty in ascertaining the true number of infections. Testing during an epidemic tends to focus on clinically severe cases, which may bias estimates of fatality rates upwards. Conversely, there is a lag time between onset of symptoms and clinical outcome, which may lead to underreporting of fatalities (Verity et al, 2020). Another reason for variation is that fatality rates depend on factors such as age, pre-existing health conditions, and availability of healthcare including ICU beds and ventilators. Using data from 37 countries, stratified by age and adjusted for under-reporting and for the lag from onset to clinical outcomes, Verity et al (Verity 2020) estimated the CFR to be 1.4% and the IFR to be 0.7%.

In this study, we estimate ethnic inequities in infection fatality rates. We combine international data on infection fatality rates (IFR) from Verity et al (Verity 2020) with demographic and health data, by ethnicity, for Aotearoa New Zealand. We use these data to produce estimates of the inequity in IFR for Māori, Pacific and NZ European by age and accounting for major comorbidities. We adjust our estimates to account for the fact that, although Māori and Pacific populations are structurally younger than other ethnic groups in New Zealand, they have shorter life expectancy and higher rates of premature death than non-Māori at all ages. Mortality rates for older Māori are shaped by their life course, which includes increased exposure to infectious disease and conditions affecting respiratory function (Yon 2014). In addition, we adjust for ethnic inequities in unmet healthcare need, which captures some of the existing structural biases and racism within the healthcare system (Waitangi Tribunal, 2019; Robson 2007). We discuss other factors, not reflected in official data, which could further increase IFR for high risk communities.

Infection fatality rate is only one aspect of the epidemiology of COVID-19 and we acknowledge that inequities between ethnicities in other factors, such as COVID-19 incidence and reduced access to healthcare services during a pandemic, could also contribute to inequities in overall health burden. We focus on IFR here because it provides a key indication of how the severity of COVID-19 could vary among ethnicities and hence help identify high-risk communities. In addition, IFR is an important input for mathematical models of COVID-19 spread and mortality. However, it will be important to refine these models to account for ethnicity-specific differences in other factors, including transmission and access to healthcare.

To date, there has been little quantitative analysis on the effects of ethnicity for COVID-19 in Aotearoa New Zealand. Given the speed at which COVID-19 can potentially spread, there is an urgent need to prepare healthcare services and to put in place measures to protect at-risk groups. To address this, we use a simplified and approximate methodology, which contains numerous limitations (see Discussion). There are also shortcomings in the data on which our estimates are based, which make it difficult to disentangle the effects of age and comorbidity. We also acknowledge the inherent problems with using IFR as a measure of fatality. As such, our results should be used as an initial guide to the potential scale of COVID-19 inequity in Aotearoa New Zealand, and the absolute values of the estimated IFRs should be viewed through the lens of Verity et al (Verity 2020).

## Methods

We obtained data on the age structure (StatsNZ Census 2018, usual resident population) of three ethnicity groups in Aotearoa New Zealand: (1) Māori, (2) Pacific, (3) NZ European/other (StatsNZ, 2020). We obtained data on life expectancy for Māori, Pacific and non-Māori (StatsNZ Infoshare, 2020) and used the non-Māori data for the NZ European/other group. International data on age-specific infection fatality rate (IFR) were obtained from Verity et al (2020). See Tables 1-2.

**Table 1.** International data on age-specific IFR (central estimate of Verity et al 2020) and age distribution of Māori Pacific and New Zealand European/other ethnicity groups in New Zealand (StatsNZ, 2020).

| Age group | 0-9 | 10-19 | 20-29 | 30-39 | 40-49 | 50-59 | 60-69 | 70-79 | 80+ |
| --- | --- | --- | --- | --- | --- | --- | --- | --- | --- |
| IFR | 0.0016% | 0.007% | 0.031% | 0.084% | 0.16% | 0.60% | 1.90% | 4.30% | 7.80% |
| Age distribution |  |  |  |  |  |  |  |  |  |
| Māori | 21.79% | 19.44% | 15.73% | 11.66% | 11.42% | 10.18% | 6.19% | 2.69% | 0.90% |
| Pacific | 23.00% | 20.60% | 17.16% | 12.14% | 10.51% | 8.46% | 4.94% | 2.33% | 0.85% |
| NZ Euro | 12.59% | 12.51% | 12.39% | 11.42% | 13.07% | 13.80% | 11.63% | 8.07% | 4.53% |

**Table 2.** Life expectancy at birth (in years) of Māori, Pacific and non-Māori ethnicities (StatsNZ, 2020).

|  | Female | Male | Average |
| --- | --- | --- | --- |
| Māori | 77.1 | 73.0 | 75.1 |
| Pacific | 78.7 | 74.5 | 76.6 |
| Non-Māori | 83.9 | 80.3 | 82.1 |

### Adjusting for life expectancy

Māori typically experience adverse health outcomes at an earlier age than non-Māori (Ministry of Health NZ, 2019b). To reflect this, we adjusted the age-specific IFR estimates of Verity et al (2020) by the most recent (2012-14) estimates of life expectancy for each ethnicity (StatsNZ 2020). This approach is consistent with international evidence that COVID-19 mortality is approximately proportional to total mortality, meaning that COVID-19 amplifies existing risk of mortality evenly for different groups (Spiegelhalter 2020). The gap in life expectancy is different for male and female, and for different age groups. However, for simplicity, we used an average of the male and female life expectancy gap for the youngest age cohort. For example, average life expectancy for Māori is approximately 8.6% less than for non-Māori. We allowed for this by assuming that, all else being equal, Māori of age *a* experience the same IFR as non-Māori who are 8.6% older. We calculated the IFR for age group *A*, adjusted for the life expectancy of ethnicity group *j*, as follows:

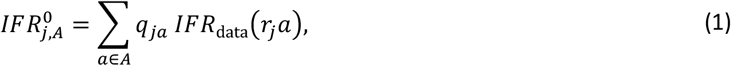

where *q*_*ja*_ is the proportion of ethnicity group *j* within age group *A* that is age *a, IFR*_data_(*a*) is the IFR at age *a* in the reference population (in which the IFR data were measured), and *r*_*j*_ is the ratio of the life expectancy of the reference population to the life expectancy of group *j*. We used 20-year age bands to reflect the breakdown of the New Zealand health data, but the distribution of ages within each age group for each ethnicity is accounted for in Eq. (1). *IFR*_data_ was evaluated at ages *r*_*j*_ *a* by linearly interpolating the data between the mid- points of the age brackets in Verity et al (2020). Age 85 was used as the mid-point for the 80+ age group, with the IFR for all ages >85 fixed at this rate.

### Adjusting for unmet healthcare need

We obtained data on unmet needs for primary healthcare for each ethnicity (Ministry of Health NZ, 2019a). The proportion of people who self-reported that they were unable to see a GP when needed (*u*_*j*_) was 41.4% for Māori, 35.9% for Pacific people and 30.1% for NZ European people. We took these data as a rough proxy for potential under-reporting of comorbid conditions and other inequities and racism within the healthcare system (see also Discussion). To account for these differences, we weighted the IFRs for each ethnicity group by these values.

### Adjusting for comorbidity

To adjust IFRs for comorbidity, we obtained data on relative CFRs from (China CDC, 2020) for four underlying health conditions known to affect COVID-19 mortality rate, broadly defined as: (1) asthma; (2) diabetes; (3) heart disease; (4) cancer. In addition, we adjusted for comorbidity associated with smoking in the same way (Guan et al, 2020). We obtained age-stratified data on the prevalence of these conditions for each of the three ethnicity groups in New Zealand (Coppell 2013; Chan 2008; Zhang 2018; Ministry of Health NZ 2019a; StatsNZ 2013) (see Table 3). The data from China CDC (2020) included data on comorbidity rates for hypertension. However, this has not been classified by New Zealand District Heath Boards as a high-risk condition (NZ Government, 2020), so we did not include hypertension in our analysis.

**Table 3.** Data on prevalence by ethnicity and age of four health conditions and smoking (Coppell et al, 2013; Chan et al, 2008; Zhang et al, 2018; Ministry of Health NZ, 2019a; StatsNZ, 2013).

| Age group | Prevalence<br>Māori | Pacific | NZ European/other |
| --- | --- | --- | --- |
| **Diabetes** |  |  |  |
| 0-19 | 0.00% | 0.00% | 0.20% |
| 20-39 | 5.50% | 10.70% | 2.80% |
| 40-59 | 20.80% | 32.90% | 6.90% |
| 60-79 | 34.70% | 34.20% | 13.20% |
| 80+ | 40.10% | 55.80% | 20.30% |
| **Heart Disease** |  |  |  |
| 0-19 | 0.00% | 0.00% | 0.00% |
| 20-39 | 1.17% | 1.10% | 0.52% |
| 40-59 | 7.46% | 6.99% | 3.78% |
| 60-79 | 25.12% | 22.26% | 17.06% |
| 80+ | 46.80% | 38.68% | 40.75% |
| **Asthma (medicated)** |  |  |  |
| 0-19 | 17.80% | 15.80% | 14.80% |
| 20-39 | 16.40% | 11.60% | 12.70% |
| 40-59 | 16.40% | 11.60% | 12.70% |
| 60-79 | 16.40% | 11.60% | 12.70% |
| 80+ | 16.40% | 11.60% | 12.70% |
| **Cancer** |  |  |  |
| 0-19 | 0.01% | 0.02% | 0.02% |
| 20-39 | 0.08% | 0.10% | 0.09% |
| 40-59 | 0.58% | 0.54% | 0.50% |
| 60-79 | 1.97% | 1.76% | 1.69% |
| 80+ | 3.15% | 2.19% | 2.78% |
| **Smoking** |  |  |  |
| 0-19 | 4.83% | 3.14% | 2.34% |
| 20-39 | 39.47% | 29.71% | 19.52% |
| 40-59 | 34.99% | 24.12% | 15.51% |
| 60-79 | 19.01% | 12.98% | 8.49% |
| 80+ | 12.18% | 7.71% | 6.52% |

To produce relative risk factors for comorbidities, we calculated the relative increase in the CFR (for the four health conditions) and the relative increase in incidence of severe cases (for smoking) by:

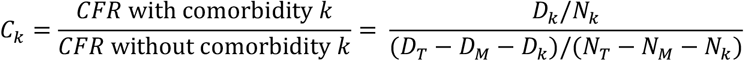

where *D*_*k*_ is the number of deaths in patients with comorbidity *k, N*_*k*_ is the number patients with comorbidity *k*. Subscripts *T* and *M* respectively represent the same quantities for the total sample and for those with missing data. Data was unavailable on the effect of smoking on CFR directly, so the incidence of severe cases was used as a proxy. The data used and relative risk factors calculated are presented in Table 4.

**Table 4.**
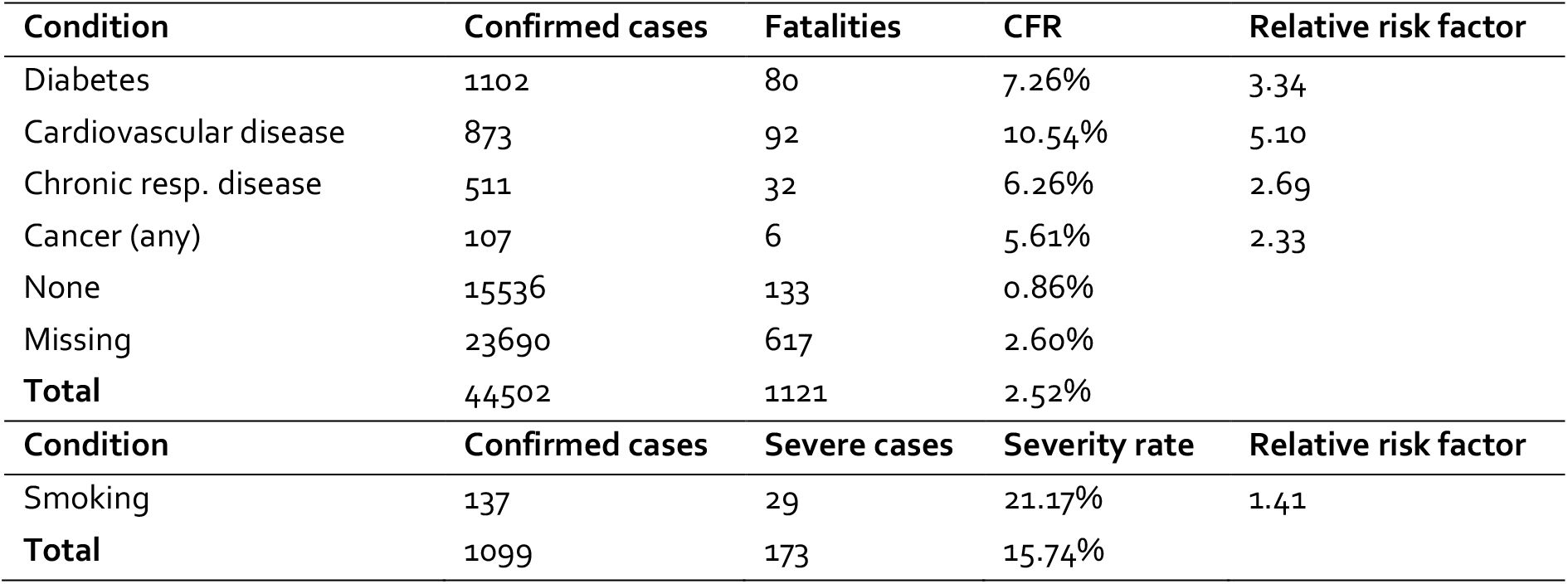
Data on fatality rates for four comorbidities obtained from (China CDC, 2020), and the effect of smoking on case severity obtained from Guan et al (2020).

| Condition | Confirmed cases | Fatalities | CFR | Relative risk factor |
| --- | --- | --- | --- | --- |
| Diabetes | 1102 | 80 | 7.26% | 3.34 |
| Cardiovascular disease | 873 | 92 | 10.54% | 5.10 |
| Chronic resp. disease | 511 | 32 | 6.26% | 2.69 |
| Cancer (any) | 107 | 6 | 5.61% | 2.33 |
| None | 15536 | 133 | 0.86% |  |
| Missing | 23690 | 617 | 2.60% |  |
| **Total** | **44502** | **1121** | **2.52%** |  |

| Condition | Confirmed cases | Severe cases | Severity rate | Relative risk factor |
| --- | --- | --- | --- | --- |
| Smoking | 137 | 29 | 21.17% | 1.41 |
| **Total** | **1099** | **173** | **15.74%** |  |

The prevalence data was reported in various age groupings; we standardised these to 20-year age groups by making the following approximation. In cases where the data were more finely stratified (cancer, smoking), we calculated a weighted average for the prevalence in 20-year bands. The diabetes data did not match the age groups, so the rates were assigned to the most similar age group (e.g. 25-44-year-old diabetes rates were assigned to the 20-39 age group). The asthma data only featured two groups: under 15 and 15+, the former was applied to the 0-19 age group and the latter to the others. There was no data on smoking rates for under 15- year olds so the rate was assumed to be 0. This is clearly an underestimate but is likely to have little impact as IFR for COVID-19 is very low in this age group.

To account for effect of comorbidity on the ethnicity-specific baseline IFRs, we made several simplifying assumptions:

1. The overall population IFR in New Zealand (across all ethnicities) is approximately equal to the overall average IFR estimates reported by Verity et al (2020) (see Discussion for more details).
2. Conditions are independent so P(condition 1 and condition 2) = P(condition 1)*P(condition 2).
3. Individuals with multiple conditions experience the product of the comorbidities of each condition, i.e. there are no interaction effects between conditions.
4. The relative effect of comorbidities on IFR is the same as the measured effect on CFR (China CDC, 2020) and is not age specific.

This allowed us to define a comorbidity weighting factor for ethnicity group *j*and age group *A* as:

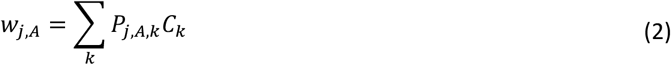

where *P*_*j,A,k*_ is the proportion of ethnicity *j* and age *A* with condition *k*, and *C*_*k*_ is the relative is the relative risk factor factor for condition *k*.

Accounting for the combined effects of age structure and comorbidity is not straightforward, as we only had data on the overall effect of each comorbidity, rather than age-specific effects. Prevalence of age-correlated conditions such as heart disease will be higher in groups with older populations. This is already reflected, to some extent, in the age distribution of IFR, which is much higher in older groups. Therefore, taking an age-structured IFR and adjusting for comorbidity from age-correlated conditions will result in some over-accounting of the effect of age-related health conditions. Similarly, separately adjusting for differences in life expectancy and prevalence of comorbid conditions will also result in some over-accounting. Conversely, ignoring age structure and only accounting for variations in prevalence of the selected health conditions may ignore some age-related effects, for example from conditions that are not in the dataset or age effects that are not linked to a specific health condition (see also Discussion).

We therefore calculated an IFR for each ethnicity group in two different ways: (i) starting with an age-specific baseline IFR and adjusting by ethnicity for life expectancy, unmet healthcare need, and comorbidity; and (ii) starting with the same population-wide baseline IFR and adjusting by ethnicity for life expectancy, unmet healthcare need, and comorbidity. The actual value likely lies somewhere between (i) and (ii), so this gives an indicative range for the scale of relative differences in IFR by ethnicity. For method (i), the equation used to calculate IFR for age group *A* and ethnicity group *j* was

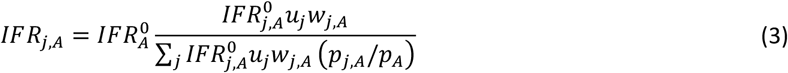

where 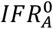 is the population average IFR of age group 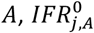 is the life-expectancy-adjusted IFR from Eq. (1), and *P*_*j,A*_ is the proportion of the population that is in age group *A* and ethnicity group *j*. The denominator of Eq. (1) normalises so that the overall population average IFR in the age group is 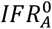, consistent with Verity et al (2020). For method (ii), the equation for the overall IFR for ethnicity *j* was

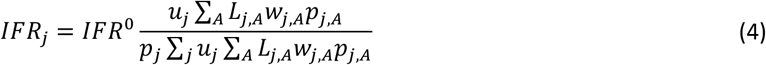

where *L*_*j,A*_ is a factor used to adjust for the effect of life expectancy on the IFR for ethnicity *j* and age group *A*. The denominator of Eq. (2) normalises so that the overall population average IFR is fixed at *IFR*^0^.

## Results

Table 5 shows estimated IFRs for each ethnicity, with and without pre-adjusting for the age structure of that ethnicity. The estimated overall population IFR is 0.81%. However, this overall rate could be influenced by numerous factors not accounted for here (see Discussion). In particular, it should be remembered that the case fatality rate (CFR) may be substantially higher due to case under-ascertainment (Verity 2020). The results shown here should be interpreted primarily as indicating the scale of relative differences in IFR across ethnicities and age groups.

**Table 5.** Estimated infection fatality rates for each ethnicity group. If age itself is the primary factor, then the results from method (i) are likely to be more accurate. If the age effect is driven by the increase in comorbidity rates with age, the results from method (ii) are likely to be more accurate.

| Method (i) | Māori | Pacific | NZ Euro./ other | Overall |
| --- | --- | --- | --- | --- |
| 0-19 years | 0.01% | 0.01% | 0.00% | 0.01% |
| 20-39 years | 0.12% | 0.09% | 0.04% | 0.06% |
| 40-59 years | 1.33% | 1.00% | 0.28% | 0.45% |
| 60-79 years | 7.88% | 5.52% | 2.22% | 2.78% |
| 80+ years | 13.87% | 11.75% | 6.76% | 7.14% |
| Overall | 1.15% | 0.72% | 0.75% | 0.81% |
| Method (ii) | Māori | Pacific | NZ Euro./ other | Overall |
| Overall | 1.66% | 1.17% | 0.62% | 0.81% |

Under both method (i) and (ii), Māori have the highest estimated IFR. Under method (i) (IFR pre-adjusted for age structure), the estimated IFR for Māori is 1.15%, compared to 0.75% for NZ European/other and 0.72% for Pacific people. Under method (ii), the disparity is larger and the estimated IFR for Māori (1.66%) more than 2.5 times that for NZ European/other (0.62%). The estimated IFR for Pacific people (1.17%) is also significantly higher than for NZ European/other. This is due to a combination of relatively low life expectancy, high unmet healthcare need, and high prevalence of underlying health conditions, notably diabetes, heart disease and asthma. The disparity is smaller under method (i) than under method (ii) because method (i) accounts for the fact that the Māori and Pacific populations are structurally younger than the NZ European/other population.

Sensitivity analysis was performed on two model assumptions: the magnitude of the difference in age-specific health outcomes between Māori/Pasifika and non-Māori; and the magnitude of the disparity in unmet healthcare need. The estimates we have used for these effects are based on indirect or proxy data (life expectancy and GP access respectively) and these are likely to be underestimates. Three scenarios are presented in Table 6: (1) the impact of the difference in life expectancy (*r*_*j*_ in Eq. (1)) between NZ European and Māori/Pasifika is doubled from 8.6% to 17.2%; (2) the discrepancy in unmet healthcare need *u*_*j*_ between NZ European and Māori/Pasifika is doubled; and (3) both adjustments. Results of these scenarios are shown in Table 6. These are shown for method (i) using age-specific IFRs.

**Table 6.** Results of sensitivity analysis of the estimated infection fatality rates on assumptions about inequities in healthcare outcomes at a given age. IFRs are pre-adjusted for age (method (i)). Darker colours indicate higher rates. For scenario (1), the change in impact of life expectancy is assumed to redistribute the rates without changing the overall IFR. In scenario (2) and (3), the increase in unmet healthcare needs is assumed to increase the overall IFR.

| Scenario |  |  |  |  |
| --- | --- | --- | --- | --- |
| **(1) Increase in impact of differences in life expectancy** |  |  |  |  |
|  | Māori | Pacific | Other | Overall |
| 0-19 years | 0.01% | 0.01% | 0.00% | 0.01% |
| 20-39 years | 0.14% | 0.10% | 0.04% | 0.06% |
| 40-59 years | 1.71% | 1.15% | 0.21% | 0.45% |
| 60-79 years | 10.12% | 6.77% | 1.98% | 2.78% |
| 80+ years | 14.16% | 12.01% | 6.74% | 7.14% |
| Total Population | 1.44% | 0.84% | 0.69% | 0.81% |
| **(2) Increase in impact of unmet healthcare need** |  |  |  |  |
|  | Māori | Pacific | Other | Overall |
| 0-19 years | 0.02% | 0.01% | 0.00% | 0.01% |
| 20-39 years | 0.24% | 0.18% | 0.04% | 0.08% |
| 40-59 years | 2.66% | 2.01% | 0.28% | 0.67% |
| 60-79 years | 15.76% | 11.05% | 2.22% | 3.59% |
| 80+ years | 27.74% | 23.51% | 6.76% | 7.91% |
| Total Population | 2.30% | 1.44% | 0.75% | 1.03% |
| **(3) Both of the above combined** |  |  |  |  |
|  | Māori | Pacific | Other | Overall |
| 0-19 years | 0.02% | 0.01% | 0.00% | 0.01% |
| 20-39 years | 0.27% | 0.19% | 0.04% | 0.09% |
| 40-59 years | 3.41% | 2.30% | 0.21% | 0.73% |
| 60-79 years | 20.25% | 13.54% | 1.98% | 3.81% |
| 80+ years | 28.33% | 24.01% | 6.74% | 7.93% |
| Total Population | 2.87% | 1.69% | 0.69% | 1.08% |

## Discussion

Disentangling the effects of age structure and comorbidity on COVID-19 infection fatality rates is difficult because international literature to date is mostly limited to univariate analysis. Also, estimates from China of the impacts of comorbid conditions are not stratified by age. Furthermore, the list of health conditions impacting on COVID-19 infections continues to be updated as the pandemic develops, with additional conditions being added. The current modelling has been limited to the list of conditions included in the study from China CDC (2020). The data from which the baseline IFRs are calculated are adjusted for under-reporting, bias towards testing more severe cases, and lag time from onset to clinical outcome (Verity 2020). Nevertheless, there may be other biases and sources of inaccuracy not accounted for. We assumed in our model that the overall IFR was fixed at 0.87%, but there may be country-specific variations in IFR, and potentially higher IFRs in countries with large ethnic minority or Indigenous populations. Therefore, the IFR results discussed here should be treated as a preliminary estimate of relative differences between ethnicities, rather than predictions of the absolute IFR.

We calculated IFRs for each ethnicity group using two different methods using the currently available New Zealand data on four underlying health conditions and smoking. This gives an approximate range of possibilities for the IFR for each group. The methodology we have used makes relatively crude adjustments to IFRs based on differences in life expectancy, unmet primary healthcare need, and prevalence of comorbid conditions. This methodology should be refined over time, particularly as more data become available on COVID-19 cases in New Zealand. An alternative approach would be to use standardised metrics such as disability-adjusted life year (DALY) and years lost due to disability (YLD) to infer IFRs by age and ethnicity in New Zealand from the Chinese data. This approach should be tested, however it is possible that the true magnitude of inequities are not captured in these metrics and the data on which they are based, so there is a risk that this approach will underestimate the health burden for Māori and Pasifika.

**Figure 1.**
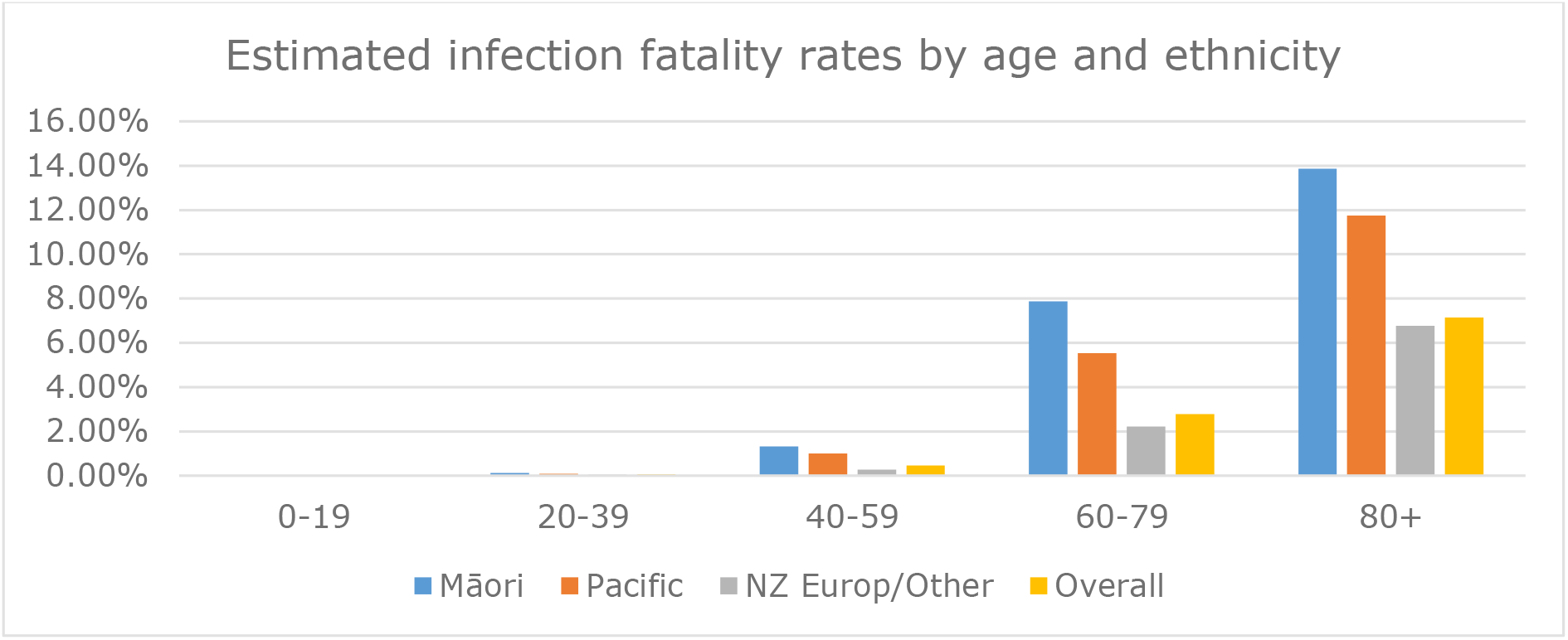
Estimated infection fatality rates by age and ethnicity using method (i). These estimates are adjusted for age structure, relative life expectancy, unmet healthcare need, and comorbidity (first section of Table 5).

The NZ European/other group have a relatively old population, but relatively high life expectancy and low unmet healthcare need. Māori and Pasifika have younger populations, but lower life expectancy, higher unmet healthcare need and higher prevalence of relevant underlying conditions such as diabetes and asthma. These factors have opposing effects on the IFR and it is difficult to predict whether age, or other covarying factors, will end up being more important. There is, as yet, little direct evidence to distinguish these effects for COVID- 19. However, regardless of whether age or co-morbidities are the dominant factor influencing IFR, Māori will be worse off than NZ European/other. If age is the dominant variable, we estimate that the IFR for Māori will be about 50% higher than that for non-Māori. On the other hand, if underlying health conditions (which correlate with age) are more important, Māori could experience an IFR more than 2.5 times higher than that of NZ European, and Pasifika could experience an IFR around 30% higher than NZ European (first section of Table 5). Māori are more likely to experience multimorbidity and if the effect of multiple underlying health conditions is worse than simply multiplicative as assumed here, this will increase the IFR for Māori. These disparities could be wider still if differences in age-specific health outcomes and unmet healthcare need are larger than captured in official data (discussed below). Hospitalisation and fatality rates for Māori and Pasifika in the H1N1 influenza epidemic of 2009 were significantly higher than for NZ European (Verrall 2010; Wilson 2012). This also suggests that, if COVID-19 mortality patterns follow H1N1 influenza mortality patterns, the IFRs for Māori and Pasifika in Table 5 could be underestimates.

Our results account in a crude way for differences in unmet primary healthcare need across ethnic groups, measured by data on GP access. However, there exists other widely reported racism within the healthcare system (Reid 2014; Cormack 2018; Ministry of Health NZ 2019b) that is not reflected in the available data. For example, access to ICU facilities may be lower for rural populations, particularly for rural Māori. Avoidable hospitalisations are higher for Māori and Pasifika (Basu 2008; Ministry of Health NZ 2019b), reflecting broader and more complex structural disadvantage than is reflected in the data. Māori and Pasifika prevalence data on comorbid conditions (Table 3) may be influenced by underreporting, which would make their IFRs higher than calculated here. All of these factors are likely to make IFRs for Māori and Pasifika higher than the estimates presented here. Table 6 shows a scenario analysis that could account for the effects of some of these factors.

These factors may not be as important during the early stages of the COVID-19 outbreak, when the goal is elimination or containment, and when surveillance and contact tracing systems are strong. However, if rapid and out-of-control spread of COVID-19 takes over, as has happened elsewhere, it will place unprecedented stress on healthcare services. This will make access to healthcare increasingly difficult and necessitate decisions by practitioners about who gets access to care. This will almost certainly amplify existing racism in the healthcare system. For example, if triage decisions are based on existence of underlying health conditions, this will automatically disadvantage Māori further. Similar concerns about the disparate impacts of prioritisation tools have been raised elsewhere (King et al, 2020). There is a need for transparency in the risk factors and weightings used to guide decision-making about access to resources, and for independent oversight by those at-risk groups most likely to be disparately impacted by these.

The IFRs presented in Table 5 using method (i) conform to the established pattern of IFR being much higher in older groups (Verity 2020). This is especially important in specific regions or communities with a relatively old population, which is likely to be one of the biggest factors affecting hospitalisation rates, ICU demand and fatality rates. For example, rural Māori have an older age distribution than Māori as a whole. Rural areas and particularly rural Māori communities may also have higher unmet healthcare need and less access to hospital-level care, so this could be a particularly high-risk group. Reported IFRs for COVID-19 do not capture indirect impacts of COVID-19, for example deaths that are attributed to underlying conditions, such as heart disease, but were precipitated or hastened by COVID-19 infection. These indirect impacts are likely to also fall disproportionately on Māori and Pasifika peoples due to their higher prevalence of comorbid conditions.

A report from the United Kingdom suggests black, Asian and other ethnic minority groups are more at-risk from COVID-19 than white majority groups (ICNARC, 2020). These at-risk communities have higher prevalence than white British groups of underlying health conditions, and are more likely to live in overcrowded households (UK Government 2018) and multi-generational households. Early reports emerging from the United States also suggest similar trends, where African-American communities are bearing a disproportionate health burden by COVID-19, thought to be related to pre-existing health conditions (e.g. (IDPH 2020)). These minority groups also have relatively young age structure (UK Government 2018). Similar factors apply to Māori in New Zealand (McLeod 2020) and this reinforces the importance of accounting for factors specific to ethnicity groups. The methodology we have used here is a first attempt at addressing this, rather than crudely using age structure alone to estimate IFR. Data on COVID-19 incidence and outcomes stratified by ethnicity, or within the context of ethnic minority or Indigenous populations that experience disparities in health and healthcare, is currently scarce. Making robust comparisons and informing interventions to eliminate inequitable outcomes requires not only more data, but data that is accessible to decision makers in a timely fashion. This reinforces the importance of systematic, comprehensive and timely data collection in Aotearoa New Zealand in order to manage this and any future epidemics

The data and results in this report only look at the infection fatality rate, i.e. the ratio of deaths to infections, in each ethnicity group. This does not account for potential ethnic differences in transmission and infection, and therefore the proportion of each ethnicity group that becomes infected. Risk factors for heightened transmission include crowded housing, which affects around 25% of Māori and around 45% of Pacific people (Schluter 2007; StatsNZ, 2018). In addition, multi-generation households increase the risk of transmission to older groups. These compounding factors mean that Māori and Pacific peoples are at higher risk of bearing a disproportionate burden of COVID-19 sickness, hospitalisation, on-going health effects, and fatality. A comprehensive analysis of these factors is outside the scope of this work. It will be critical to incorporate these into disease transmission models that are used to inform Aotearoa New Zealand’s COVID-19 response (James 2020). This will enable the combined effect of incidence and IFR to be more accurately predicted and to inform effective strategies that recognise the diversity of higher risk groups and regions.

## Data Availability

The article does not present new primary data.

## Acknowledgements

The authors would like to thank Tony Blakely, Nigel French, Ricci Harris, Karen Wright, Thomas Lumley, and Collin Tukuitonga for helpful comments on the manuscript.

